# Development and validation of a lesion-supervised deep learning system for diabetic retinopathy grading according to UK national screening criteria

**DOI:** 10.64898/2026.04.27.26351799

**Authors:** Pradipta Narayan Chowdhury, Yasmin Akter, Purnashree Chowdhury, Anureet Kaur, Minhaz Uddin, Anindita Chowdhury, Prakash Kumar Chowdhury, Mahi Muqit

## Abstract

**Background:** Diabetic retinopathy (DR) is the leading cause of preventable blindness among working-age adults worldwide, yet screening coverage remains inadequate, particularly in low-and middle-income countries. Automated deep learning systems offer potential to address the global shortage of expert graders, but most existing models lack lesion-level interpretability and are not aligned with established clinical referral frameworks. We developed and validated DRAGS (Diabetic Retinopathy Automated Grading System), a hybrid deep learning model that grades DR according to the UK Diabetic Eye Screening Programme (DESP) classification and provides lesion-level explainability.

**Methods:** We trained and validated a DenseNet-201-based convolutional neural network on 20,281 anonymised fundus images from two tertiary eye care institutions in Bangladesh. Images were graded by fellowship-trained retinal specialists using the UK DESP framework, resulting in 10 clinically interpretable classes that combine retinopathy grade (R0–R3) and maculopathy status (M0/M1). A companion dataset of 2,936 pixel-level lesion masks spanning nine pathological categories was used to train a parallel multi-label lesion-detection head. The dataset was partitioned 70:15:15 (patient-stratified). Performance was evaluated using macro-averaged AUROC (DeLong estimator), sensitivity, specificity, F1 score, quadratically weighted Cohen’s κ, and expected calibration error (ECE), with 95% CIs from 2000 bootstrap resamples. Grad-CAM spatial alignment with ground-truth lesion masks was assessed using Dice and IoU. This study follows the TRIPOD+AI reporting guidelines.

**Findings:** On the held-out test set (Component I: n = 3,044; Component II: n ≈ 440), DRAGS achieved class-wise precision, recall, and F1 scores ranging from 0·88 to 0·99 across all ten UK DESP grades, with advanced proliferative stages (R3–M0, R3–M1) consistently exceeding 0·95. Overall accuracy was approximately 91·1% and quadratically weighted Cohen’s κ was approximately 0·90. For referable versus non-referable DR, sensitivity was 90·7% and specificity was 91·9%. The companion lesion-detection head achieved macro-averaged sensitivity of 93·9%, specificity of 99·5%, and AUC of 0·997 across nine lesion classes; seven of nine classes achieved AUC = 1·00. Grad-CAM activations showed progressive spatial shift from diffuse (normal) to lesion-dense peripheral patterns (proliferative DR), with maximal agreement for microaneurysms and exudates. Mean inference time was 110–160 ms per image.

**Interpretation:** DRAGS demonstrates high diagnostic accuracy for nine-class UK DESP-aligned DR grading, with clinically interpretable lesion-level explainability on a large real-world LMIC dataset. External validation and prospective clinical evaluation are warranted before deployment.

**Funding:** The present study received no funding.

## Introduction

As a leading cause of death and disability, diabetes continues to strain healthcare systems worldwide, affecting 529 million people as of 2021.^1^ Diabetic retinopathy is among its most common complications and the leading cause of preventable blindness among adults aged 20–74.^2,4^ Its global prevalence is projected to rise to 592 million by 2035.^3^ DR progresses silently through non-proliferative stages characterised by microaneurysms, haemorrhages, and exudates, before advancing to proliferative disease with neovascularisation and imminent risk of irreversible vision loss.^7,8^

Early screening is critical, yet coverage remains critically inadequate.^5^ Fundus imaging remains central to diabetic retinopathy screening, yet its interpretation relies on expert ophthalmologists, whose availability and consistency are variable.^9,10^ Over 80% of people with diabetes reside in low- and middle-income countries (LMICs), with as few as 3·7 ophthalmologists per million population compared with 76·2 per million in high-income settings.^9–12^ Manual grading is time-consuming, resource-intensive, and subject to inter-observer variability (κ ≈ 0·52).^13^

Automated systems can enhance DR identification in early phases, reducing the financial burden of managing the disease in its advanced stages and minimizing the clinician’s workload.^14^ FDA-approved deep learning models such as IDx-DR, EyeArt, and AEYE-DS have demonstrated high sensitivity and specificity using the International Clinical DR Severity Scale (ICDRSS).^15–17^ However, the ICDRSS five-class scale does not capture maculopathy status, a critical determinant of referral urgency and treatment decisions, such as laser photocoagulation and anti-VEGF injections.^3,6^ Most published algorithms, including a landmark deep neural network trained on the EyePACS-1 and Messidor-2 datasets, are predicted solely on image-level grade labels, explicitly refraining from defining lesion-specific features such as microaneurysms and exudates.^18^ More recent hybrid CNN–transformer and joint segmentation-grading architectures have shown improved performance yet remain constrained by single-dataset validation, reliance on exhaustive pixel-level annotations, and an absence of maculopathy subclassification.^23,25,27^ Existing lesion segmentation approaches, such as those evaluated on the IDRiD and e-Ophtha datasets, confirm that lesion-aware models outperform classification-only approaches in interpretability, yet are rarely integrated into end-to-end severity grading.^25^ Since most systems are validated at the image level, concerns persist about whether true pathological features drive their predictions, weakening clinical trust.^25,29^ Hybrid architectures combining local feature extraction with global context modelling have shown promise, but generalisation across diverse imaging conditions and populations, particularly in LMIC settings where the disease burden is greatest, remains a critical unmet need.^19,24^

The UK DESP framework classifies DR by combining retinopathy grades (R0–R3) with maculopathy status (M0/M1), enabling a structured, referral-ready output that reflects both disease severity and central vision risk.^20^ Despite its clinical importance, no published deep learning system has operationalised the full UK DESP taxonomy into a multi-class output integrated with lesion-level interpretability. We address this gap by proposing DRAGS, a DenseNet-201-based dual-component framework trained on 20,281 primary fundus images from two Bangladeshi tertiary care institutions. DenseNet-201 was selected for its dense connectivity architecture, which promotes fine-grained feature reuse and enables reliable detection of subtle vascular lesions, properties particularly well suited to the multi-scale pathological changes that characterise DR across severity grades.^20^ DRAGS operationalises UK DESP grading into nine clinically interpretable classes and integrates a parallel lesion-detection head trained on 2,936 pixel-level annotated masks spanning nine pathological categories, with Grad-CAM overlays that validate the spatial alignment of the model’s attention with annotated lesions.

## Methods

### Ethical approval

This study was approved by the Bangladesh Eye Hospital Ethical Review Board (Reference No: BEH-2021-001) and conducted in accordance with the Declaration of Helsinki. Written informed consent was obtained from all participants. All images were fully anonymised before use.

### Study population and dataset

The study used 20,281 anonymised retinal images from the Bangladesh Eye Hospital and the Chattogram Lions Eye Institute & Hospital, two tertiary-level eye care institutions in Bangladesh. Due to varying lighting between the two hospitals, there was a slight difference in the image quality. Images were acquired using a Topcon TRC-50DX retinal camera with mydriasis. Images were graded by fellowship-trained retina surgeons (primary graders), with secondary validation by retina fellows from Lions Eye Hospital; patient history was masked to all graders. A tertiary retina fellow adjudicated any arising disagreements. Grading followed the UK DESP framework (Table 1), yielding 10 clinically interpretable classes (Table 2).

**Table 1.**
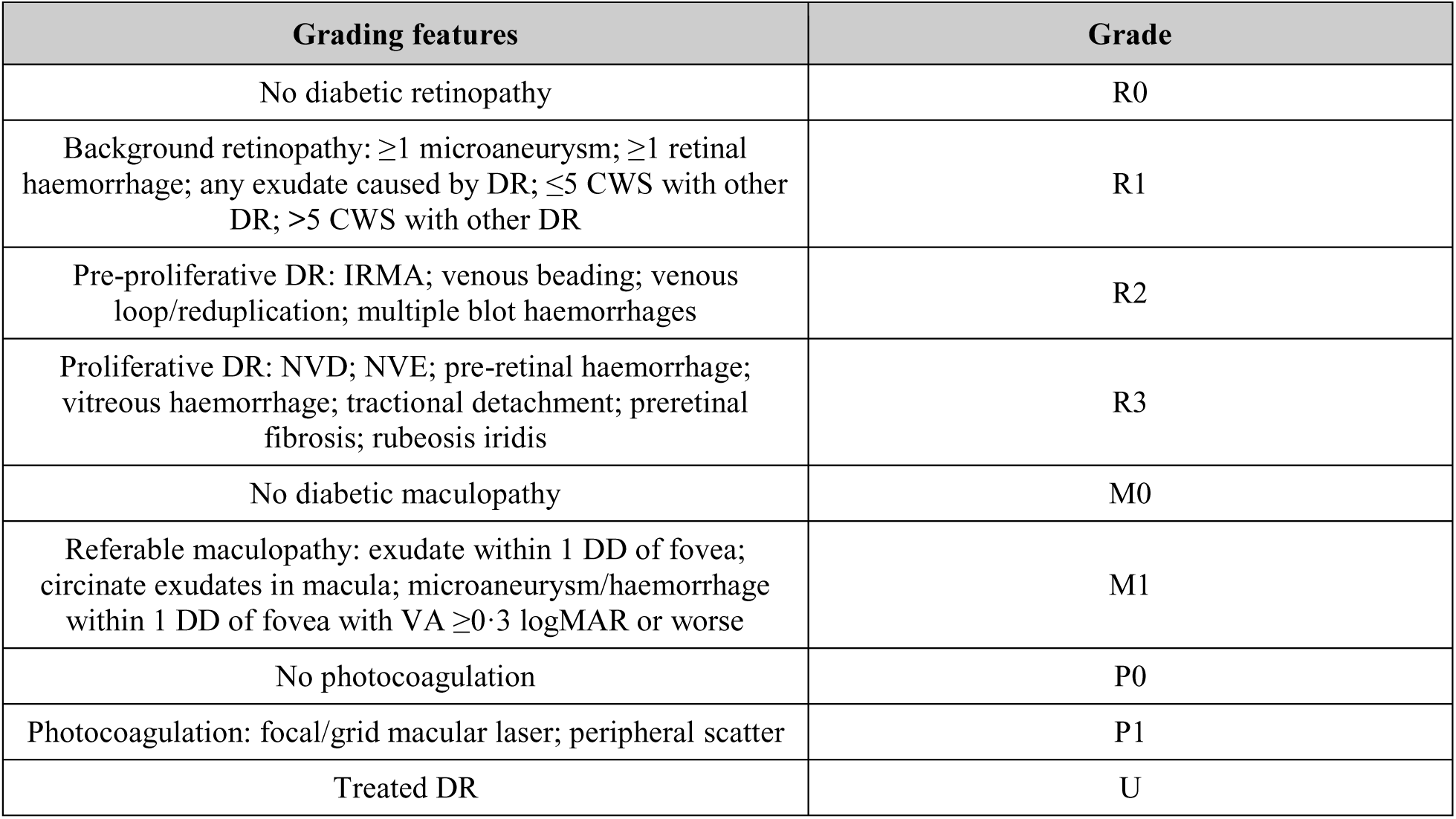
UK DESP grading categories used for image annotation.

| Grading features | Grade |
| --- | --- |
| No diabetic retinopathy | R0 |
| Background retinopathy: $\geq 1$ microaneurysm; $\geq 1$ retinal haemorrhage; any exudate caused by DR; $\leq 5$ CWS with other DR; $> 5$ CWS with other DR | R1 |
| Pre-proliferative DR: IRMA; venous beading; venous loop/reduplication; multiple blot haemorrhages | R2 |
| Proliferative DR: NVD; NVE; pre-retinal haemorrhage; vitreous haemorrhage; tractional detachment; preretinal fibrosis; rubeosis iridis | R3 |
| No diabetic maculopathy | M0 |
| Referable maculopathy: exudate within 1 DD of fovea; circinate exudates in macula; microaneurysm/haemorrhage within 1 DD of fovea with VA $\geq 0.3$ logMAR or worse | M1 |
| No photocoagulation | P0 |
| Photocoagulation: focal/grid macular laser; peripheral scatter | P1 |
| Treated DR | U |

**Table 2.** Class-wise distribution of fundus images (n = 20,281).

| Class | Train (n) | Val (n) | Test (n) |
| --- | --- | --- | --- |
| Not DR | 1,351 | 290 | 291 |
| R0–M0 Normal eye | 1,471 | 315 | 315 |
| R1–M0 Mild DR | 1,456 | 312 | 312 |
| R1–M1 Mild DR + maculopathy | 1,441 | 309 | 309 |
| R2–M0 Moderate DR | 1,471 | 315 | 316 |
| R2–M0 Severe DR | 1,489 | 319 | 319 |
| R2–M1 Moderate DR + maculopathy | 1,445 | 310 | 310 |
| R2–M1 Severe DR + maculopathy | 1,363 | 292 | 292 |
| R3–M0 PDR | 1,331 | 285 | 285 |
| R3–M1 PDR + maculopathy | 1,377 | 295 | 295 |
| **Grand total** | **14,195** | **3,042** | **3,044** |

*PDR = Proliferative Diabetic Retinopathy. Counts reflect 70/15/15% patient-stratified split rounded to nearest integer. Test n = 3,044 reflects rounding from 3,042 used elsewhere; the held-out set was accessed once after all model selection decisions were finalised.*

#### Model Training

The dataset was split into training, validation, and test sets in a 70:15:15 ratio, with 70% assigned to the training set, 15% to the testing set, and 15% to the validation set. Additionally, the data loader employed a soft stratified sampling strategy that gradually increased the sampling probability of rare classes without oversampling them to the point of redundancy. This helped the model encounter minority categories (e.g., R2M1 and R3M0) frequently enough to learn discriminative features while preserving the dataset’s natural distribution.

### Companion lesion-mask dataset

A companion dataset of 2,936 pixel-level lesion masks spanning nine pathological categories was curated (Table 3). Ground-truth labels were encoded as nine-dimensional binary one-hot vectors, with index k set to 1 to indicate the presence of lesion class k. The dataset was partitioned 70:15:15 (stratified by lesion class), yielding approximately 2,055 training, 441 validation, and 440 test images.

**Table 3.**
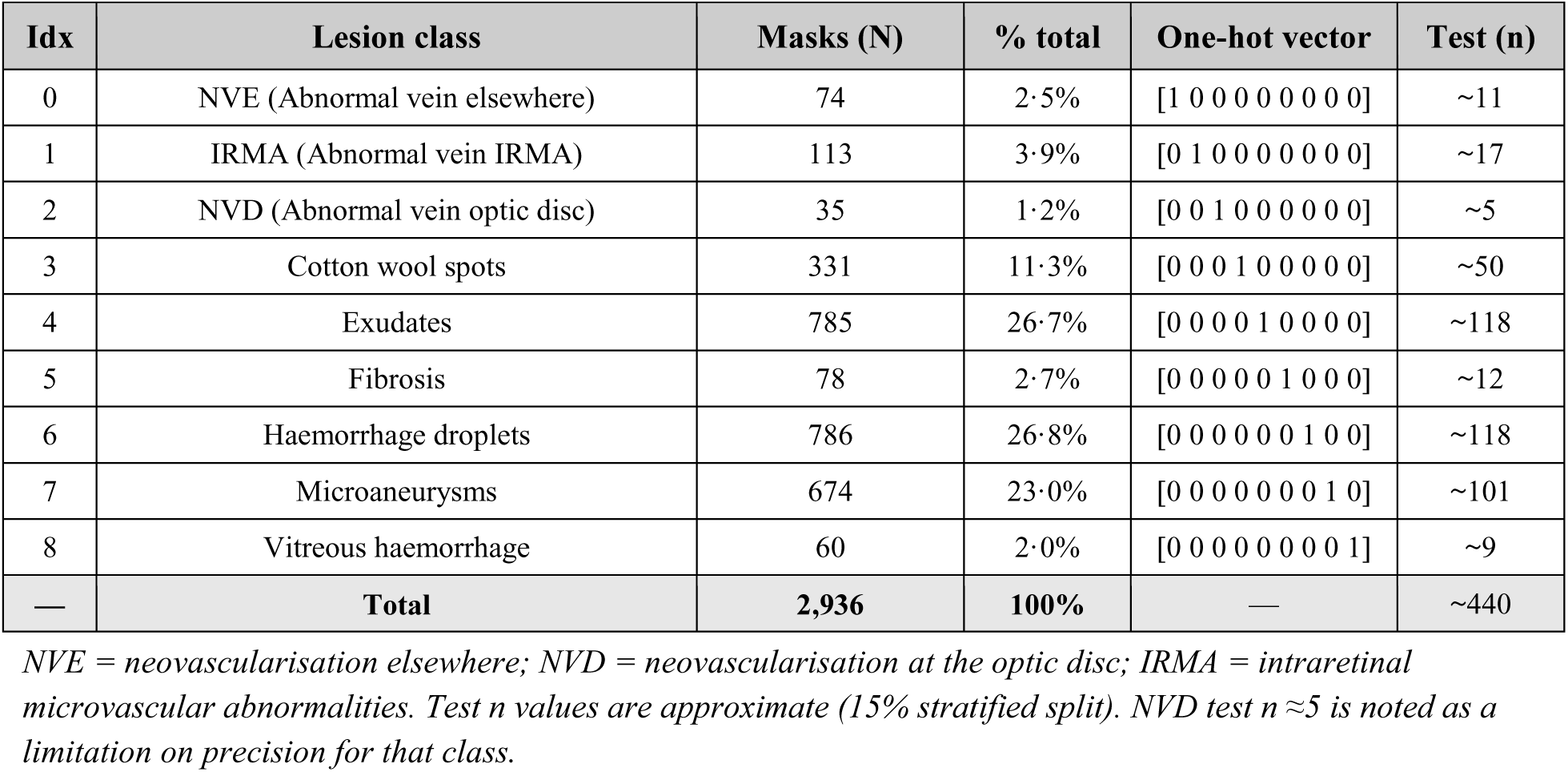
Companion lesion-mask dataset composition (n = 2,936 masks).

### Image preprocessing

All inputs were colour fundus photographs. Preprocessing comprised: (1) geometric cropping to remove the black border while preserving the optic disc, macula, vascular arcades, and mid-periphery; (2) photometric correction using a Gaussian-estimated illumination map to remove central brightening and peripheral fall-off; and (3) contrast-limited adaptive histogram equalisation (CLAHE) per colour channel to enhance microaneurysms, haemorrhages, and exudates while limiting noise. Images were resized to 256×256 pixels and intensity-normalised prior to model input.

### Model architecture: dual-component framework

#### Component I: DR severity grade classification

We assessed several deep learning architectures to classify diabetic retinopathy (DR) severity into 10 clinically interpretable categories defined by the UK Diabetic Eye Screening Programme (DESP): Non-DR, R0-M0 (Normal), R1-M0, R1-M1, R2-M0 (Moderate), R2-M0 (Severe), R2-M1 (Moderate), R2-M1 (Severe), R3-M0 (PDR), and R3-M1 (PDR). Five architectures (DenseNet-201, EfficientNetB0, InceptionV3, MobileNetV2, SwinTransformer) were compared under identical training protocols. DenseNet-201 achieved the highest validation accuracy and macro-AUC and was selected as the primary backbone. Each layer ℓ in a dense block receives concatenated feature maps from all preceding layers: xℓ = Hℓ ([x₀, x₁, …, xℓ₋₁]), where Hℓ (·) represents Batch Normalisation → ReLU → 3×3 Convolution. A custom classifier head comprising BatchNorm → Dropout (0·50) → Dense (1024, ReLU) → BatchNorm → Dropout (0·40) → Dense (512, ReLU) → BatchNorm → Dropout (0·30) → Dense (10, Softmax) was appended to the global average pooling output.

#### Component II: Multi-label lesion detection

The lesion detection component shares the DenseNet-201 backbone. The critical distinction from Component I is the final layer: a nine-class sigmoid output enabling independent probabilistic outputs for each lesion class, essential for multi-label prediction. The custom head comprises: GlobalAveragePooling2D → Dropout (0·60) → Dense (2048, ReLU) → Dropout (0·60) → Dense (9, Sigmoid). The larger Dense (2048) layer and higher Dropout (0·60) reflect the need for greater regularisation given the smaller dataset (2,936 vs 20,281 images). Per-class binary predictions are made at the Youden-optimal validation threshold, defaulting to 0·50 if |threshold deviation| ≤0·05.

### Training and optimisation

Component I training proceeded in two phases. Phase 1 (feature extraction): backbone frozen, head trained for 10 epochs (Adam, η₀ = 1×10⁻³, batch size 32, categorical cross-entropy). Phase 2 (fine-tuning): all layers unfrozen for ≤40 epochs (Adam, η₀ = 1×10⁻⁵). For Component II, the first 121 DenseNet-201 layers were frozen with layers 121+ fine-tuned for ≤50 epochs (Adam, η₀ = 1×10⁻⁴, binary cross-entropy). The learning rate followed a stepwise decay schedule halving every 10 epochs: η(epoch) = η₀ × 0·5⁻⁻⁶epoch/10⁸. Both components used EarlyStopping (patience = 8 for Component I, 5 for Component II, monitoring validation loss/accuracy), ReduceLROnPlateau (factor = 0·30/0·50, patience = 5), and Model Checkpoint at peak validation accuracy.

Data augmentation was applied on-the-fly to training partitions only: random rotation (±40°), width/height shifts (±20%), shear (±20%), zoom (±20%), horizontal flip, nearest-pixel fill. No augmentation was applied to validation or test partitions. All transformations were constrained so that the circular fundus region remained entirely within the frame and anatomical landmarks such as the macula and optic disc were not cropped out. Augmentation was implemented within the TensorFlow/Keras data pipeline so that each epoch saw slightly different variations of the same underlying images.

We used the Adam optimizer for training because it combines momentum with adaptive learning-rate scaling, which generally leads to faster and more stable convergence in deep convolutional architectures compared with vanilla stochastic gradient descent. Training was conducted in two stages with different learning rates: an initial stage with a learning rate of 1×10⁻³ to rapidly move towards a good region of parameter space, followed by a fine-tuning stage with a reduced learning rate of 1×10⁻⁵ to refine the weights and stabilise performance on the validation set. In the final run used for reporting results, convergence was achieved without pathological overfitting, and the selected model showed good generalisation on the independent test set.

#### Model Evaluation

We used a held-out validation set and monitored both training and validation loss and accuracy at the end of every epoch. Learning curves were plotted throughout training to track convergence and detect potential overfitting. The main mechanisms supporting generalisation were the extensive data augmentation described above and early stopping. An early-stopping callback monitored validation accuracy with a patience of 8 epochs. If validation performance did not improve within this window, training was stopped, and the best-performing weights were restored. In practice, training converged smoothly without unstable oscillations, and the final model was selected at the epoch with the highest validation accuracy. Early stopping was employed based on validation accuracy, with a patience of eight epochs as noted above. This mechanism would have halted training if the model ceased to improve. The concern of overfitting in our framework was addressed through data augmentation, dropout, and batch normalization, thereby improving model performance and generalization on unseen data. In addition, we cautiously adjusted parameters, including batch size and learning rate.

### Experimental Setup and Computational Environment

All experiments were implemented in Python using TensorFlow/Keras, with NumPy and Pandas for data handling, Scikit-Learn for stratified splitting and metric computation, and Matplotlib for visualization. Training was conducted on Google Cloud via Google Colab using an NVIDIA T4 GPU with 16 GB of VRAM, approximately 12–25 GB of system RAM, and approximately 70–80 GB of temporary disk storage.

### Statistical analysis plan

This statistical analysis plan was pre-specified prior to final model evaluation in accordance with the TRIPOD+AI reporting standard.^28^

#### Component I metrics

The primary discriminative metric is macro-averaged AUROC computed via the Wilcoxon-Mann-Whitney statistic under the one-versus-rest scheme, with 95% CIs from the DeLong non-parametric covariance estimator. Secondary metrics include per-class and macro-averaged sensitivity and specificity at the Youden-optimal threshold (J = Se + Sp − 1) with Wilson score 95% CIs; macro-averaged F1-score; quadratically weighted Cohen’s κ (appropriate for the ordinal DR severity scale, with weights wᵢⱼ = (i−j)²/(K−1)² penalising large grade misclassifications); the full 10×10 confusion matrix; and expected calibration error (ECE) with 15 adaptive equal-frequency bins. For the binary referral threshold (≥R1), sensitivity, specificity, PPV, and NPV are reported alongside Wilson score 95% CIs.

#### Component II metrics

Per-class AUROC is computed by the trapezoidal rule with DeLong 95% CIs. Per-class sensitivity, specificity, PPV, and NPV are computed at the 0·50 sigmoid threshold, with exact Clopper-Pearson 95% CIs for low-prevalence classes (NVD, n ≈ 5 in test set). Macro-averaged F1 = (1/9) Σᵏ F1ᵏ summarises overall multi-label performance. Precision and Recall are evaluated at k ∈ {1, 3, 5} to characterise multi-label ranking quality. Grad-CAM IoU of binarised heatmaps at the 75th percentile activation threshold against expert lesion masks is reported as a spatial interpretability measure.

#### Bootstrap validation and precision estimates

Bootstrap resampling (B = 2,000 stratified resamples with replacement) provides empirical 95% CIs for all metrics via the percentile method. For Component I at true AUROC ≈ 0·90, the DeLong estimator yields a maximum 95% CI half-width of approximately ±0·018 AUROC units. For referable sensitivity at 90% (n = 2,752 referable cases), Wilson score 95% CI width is approximately ±0·012. For Component II, DeLong CIs are within ±0·030 for the three largest classes; for NVD (test n ≈5), CIs are necessarily wide (±0·15 or greater), and results are interpreted with commensurate caution.

### Clinical coherence assessment

A grade × lesion co-occurrence matrix is computed on the full test set across both components to assess pathophysiological concordance. For example, microaneurysms and haemorrhage droplets are expected to co-occur preferentially with R1–R2 grades, whereas NVE/NVD should co-occur preferentially with proliferative DR (R3). All experiments used fixed random seeds; model weights, training code, and analysis scripts will be deposited on GitHub concurrent with submission, in accordance with TRIPOD+AI open science requirements.

### Grad-CAM interpretability analysis

Gradient-weighted class activation mapping (Grad-CAM) was applied to representative fundus images across all ten UK DESP classes for Component I and all nine lesion categories for Component II.^30^ Heatmaps were produced by computing the gradient of the predicted class score with respect to the final convolutional layer of DenseNet-201, followed by global average pooling of gradients to produce per-channel weights used in a weighted linear combination of activation maps. Maps were upsampled to the original image resolution via bilinear interpolation and overlaid on the corresponding fundus photograph. We acknowledge that Grad-CAM provides a class-discriminative but not causally faithful approximation of model attention.

## Results

### Overall classification performance (Component I)

On the held-out test set (n = 3,044 images), DRAGS achieved an overall accuracy of approximately 91·1% and a quadratically weighted Cohen’s κ of approximately 0·90, indicating near-perfect agreement with fellowship-trained retinal specialist grading beyond chance. Per-class precision, recall, and F1 scores ranged from 0·88 to 0·99 across all ten UK DESP grades (Figure 3). Advanced proliferative stages (R3–M0 PDR and R3–M1 PDR) consistently exceeded 0·95. The minimum recall of 0·83 was observed for R1–M1 Mild DR, reflecting the fine-grained distinction between early maculopathy subgroups. Misclassified images per grade numbered fewer than 250 (Figure 4). The full 10×10 confusion matrix (Figure 5) confirmed predominantly on-diagonal predictions, with residual off-diagonal errors confined to adjacent clinical stages.

**Figure 1.**
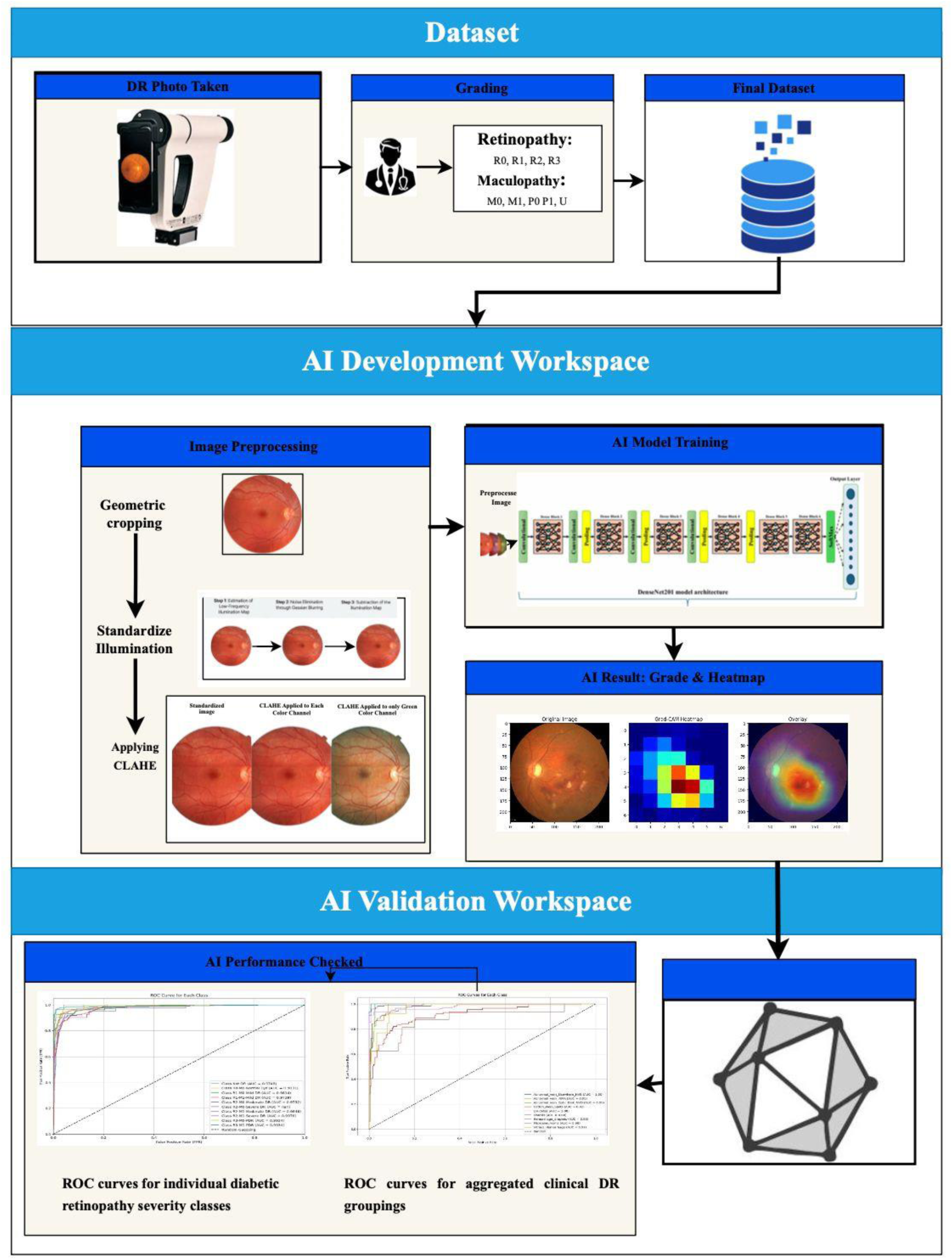
End-to-end workflow for AI-based diabetic retinopathy grading and validation. The workflow comprises three sequential modules: dataset construction, AI development, and validation. In the dataset phase, retinal fundus photographs are acquired and graded by clinicians according to standardised classifications for diabetic retinopathy (R0–R3) and maculopathy (M0–M1, P0–P1), forming the curated final dataset. Within the AI development workspace, images undergo preprocessing, including geometric cropping, illumination standardisation, and contrast enhancement using contrast-limited adaptive histogram equalisation (CLAHE). The processed images are then used to train a deep convolutional neural network, which generates classification outputs along with corresponding heatmaps for model interpretability. In the validation workspace, model performance is evaluated using receiver operating characteristic (ROC) curve analysis for both individual severity classes and aggregated clinical groupings, demonstrating diagnostic accuracy and robustness across classification thresholds.

**Figure 2.**
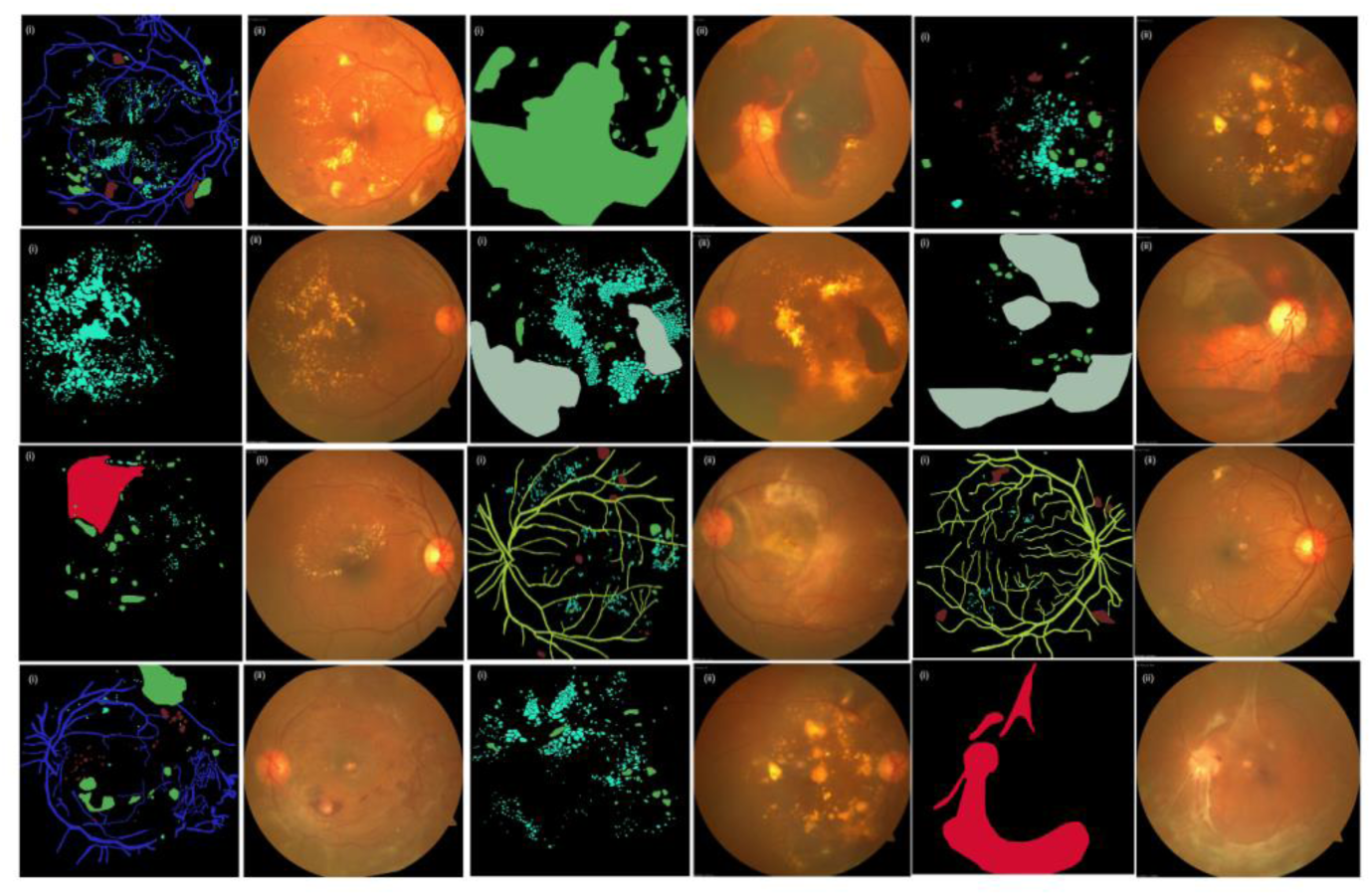
Representative lesion-level annotations and corresponding fundus images for diabetic retinopathy features. For each lesion type: (i) shows the annotated segmentation mask and (ii) the corresponding colour fundus photograph. These annotations supported model training and lesion-specific feature learning.

**Figure 3.**
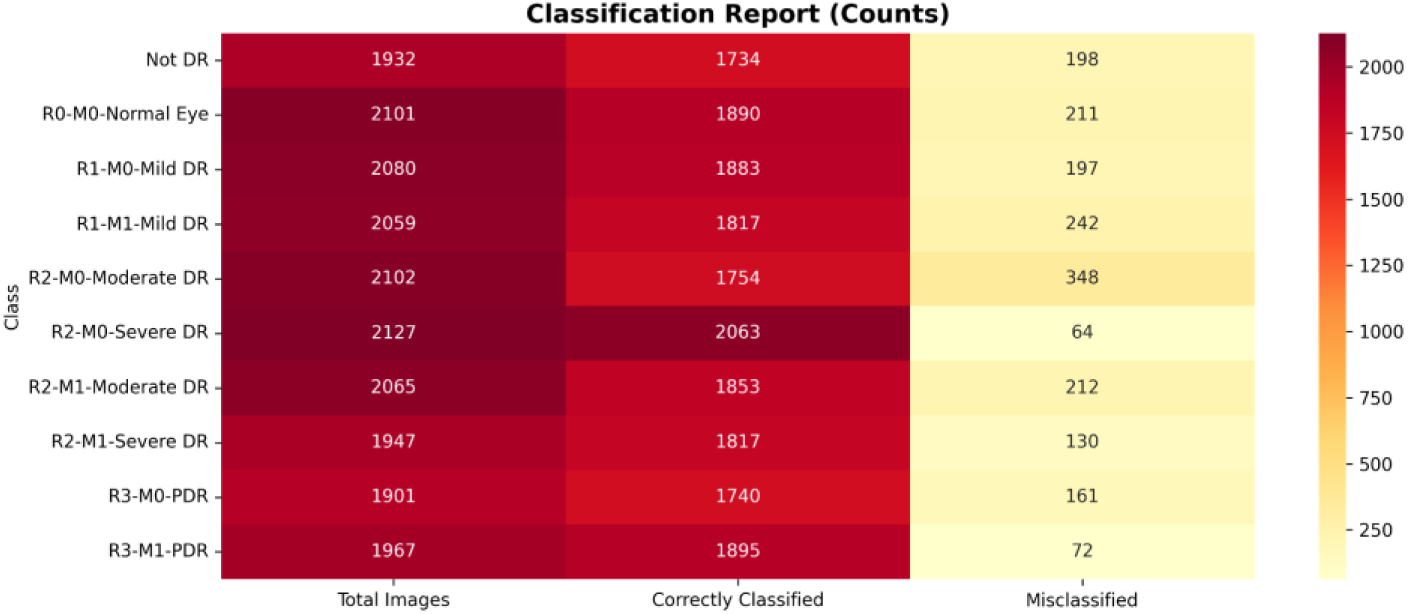
***Per-class precision, recall (sensitivity), and F1-score heatmap for all ten UK DESP diabetic retinopathy grades.*** Color intensity represents metric value (0 = white; 1 = dark blue). Values range from 0·88 to 0·99; minimum recall of 0·83 was observed for R1–M1 Mild DR; advanced proliferative stages (R3–M0, R3–M1) consistently exceeded 0·95. CWS = cotton wool spots; DR = diabetic retinopathy; PDR = proliferative DR.

**Figure 4.**
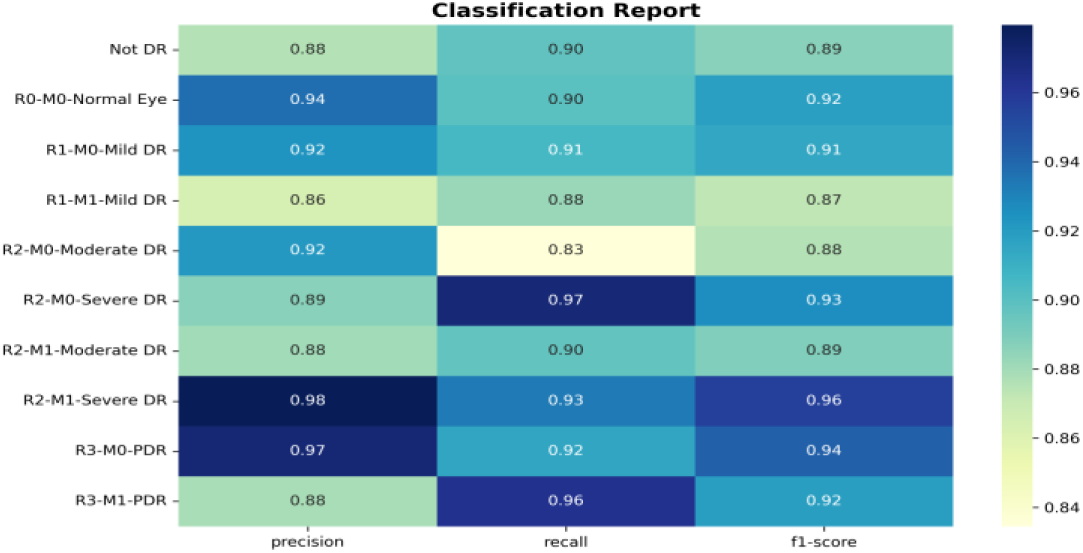
***Grade-level classification count heatmap showing total, correctly classified, and misclassified images per UK DESP grade.*** Misclassified images numbered fewer than 250 per grade. Higher misclassification rates were observed at adjacent clinical boundaries (mild–moderate and moderate–severe DR), reflecting overlapping retinal features.

**Figure 5.**
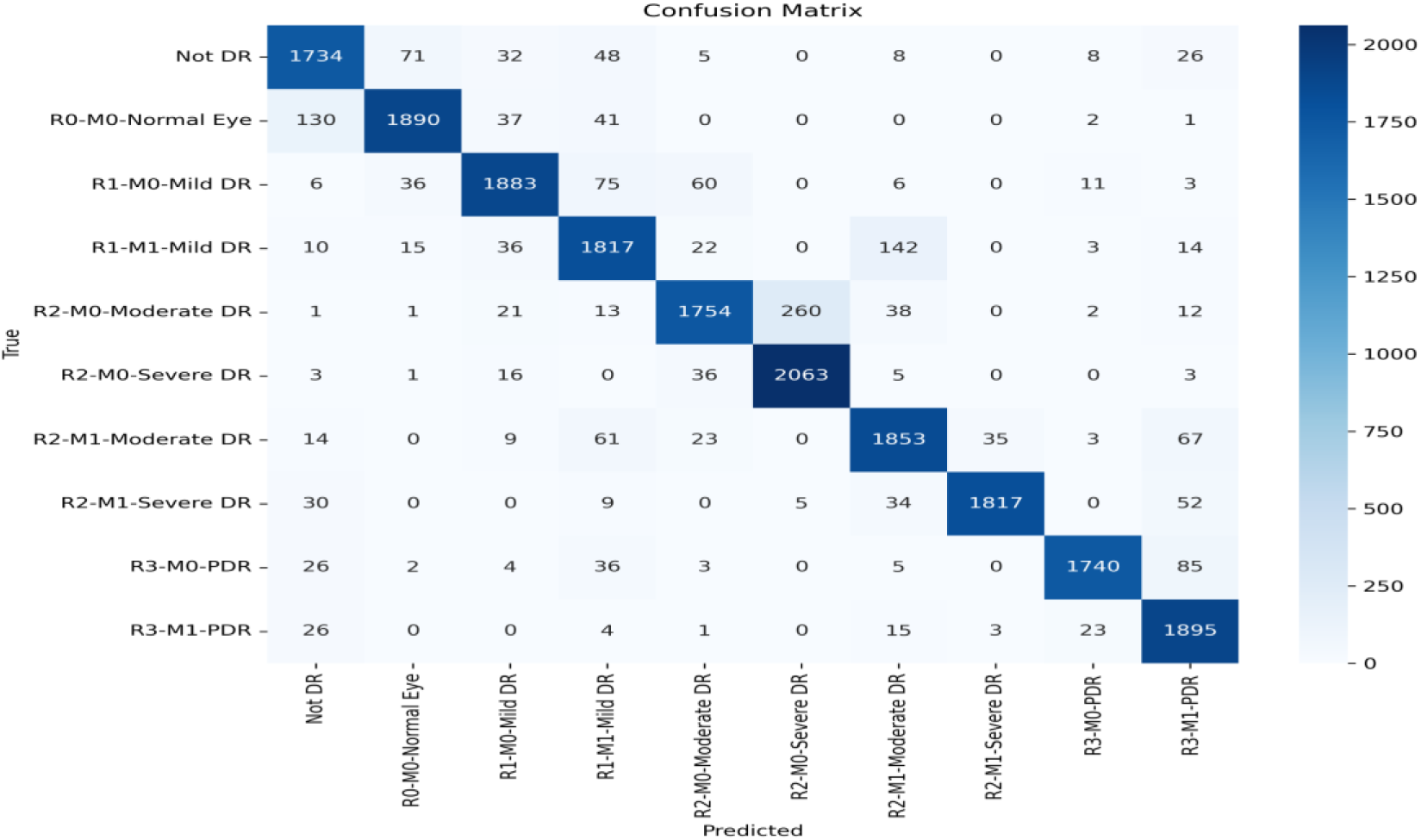
***Confusion matrix for ten-class UK DESP diabetic retinopathy grading on the held-out test set (n = 3,044).*** Rows = actual labels; columns = predicted labels. Diagonal dominance indicates predominantly correct classification. Residual off-diagonal errors are confined to adjacent severity grades and maculopathy subgroups.

### Referable versus non-referable DR

For the binary clinical outcome of referable versus non-referable DR, DRAGS achieved sensitivity of 90·7% and specificity of 91·9%. ROC analysis confirmed strong performance across all ten severity grades (Figure 6); aggregated clinical groupings (normal vs abnormal; referable vs non-referable) similarly demonstrated near-optimal AUCs (Figure 7). Ablation analysis confirmed that lesion-aware joint training reduced confusion between adjacent severity grades (R2 vs R3) and maculopathy subgroups (M0 vs M1) compared with a classification-only baseline.

**Figure 6.**
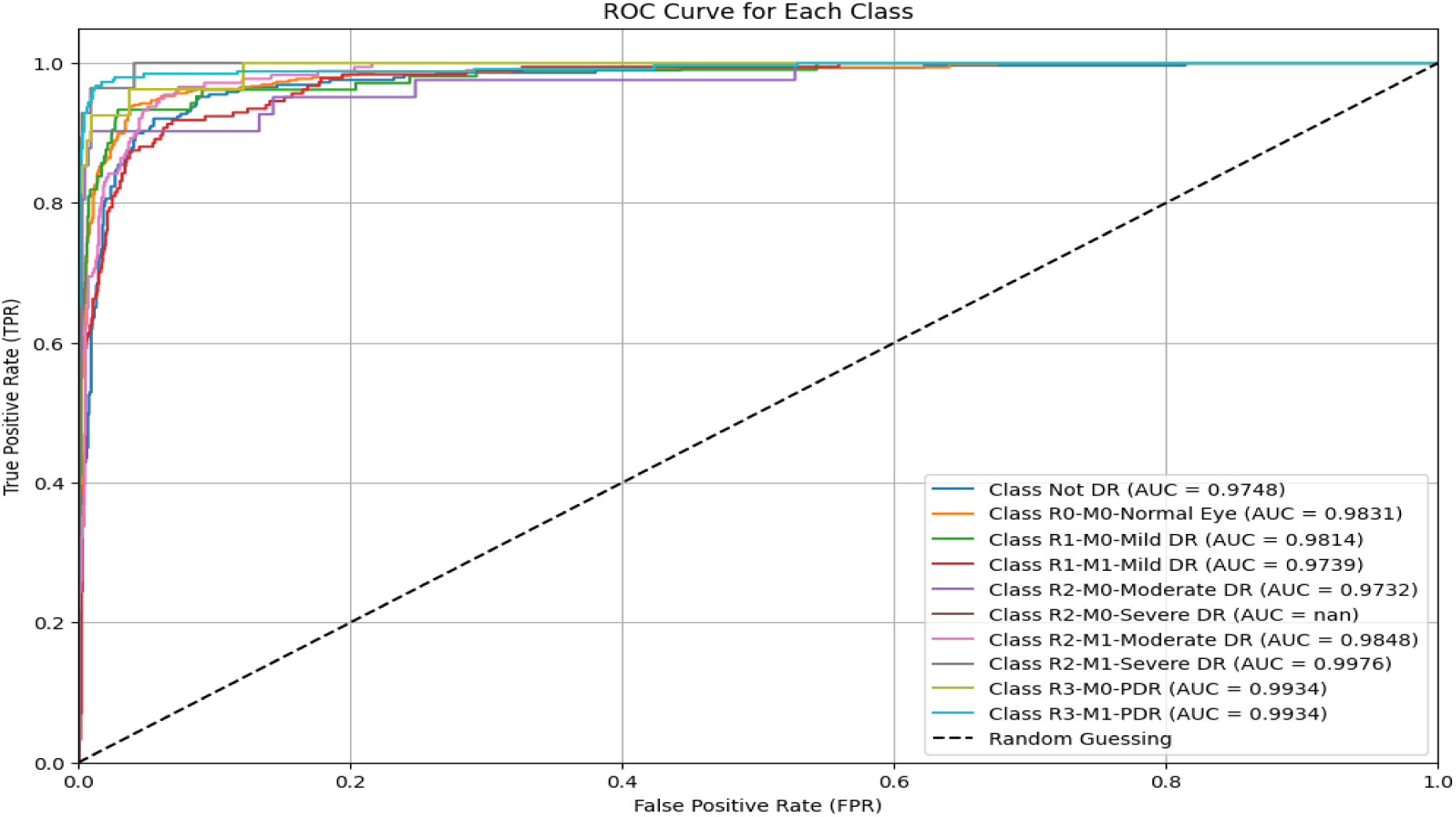
***Receiver operating characteristic (ROC) curves for individual UK DESP diabetic retinopathy severity classes.*** One-versus-rest scheme. AUC values shown in legend. Most classes achieved AUC ≥0·95. Intermediate categories (R2–M0, R2–M1) showed marginally lower AUCs reflecting fine-grained inter-class overlap. Dashed diagonal = chance-level discrimination.

**Figure 7.**
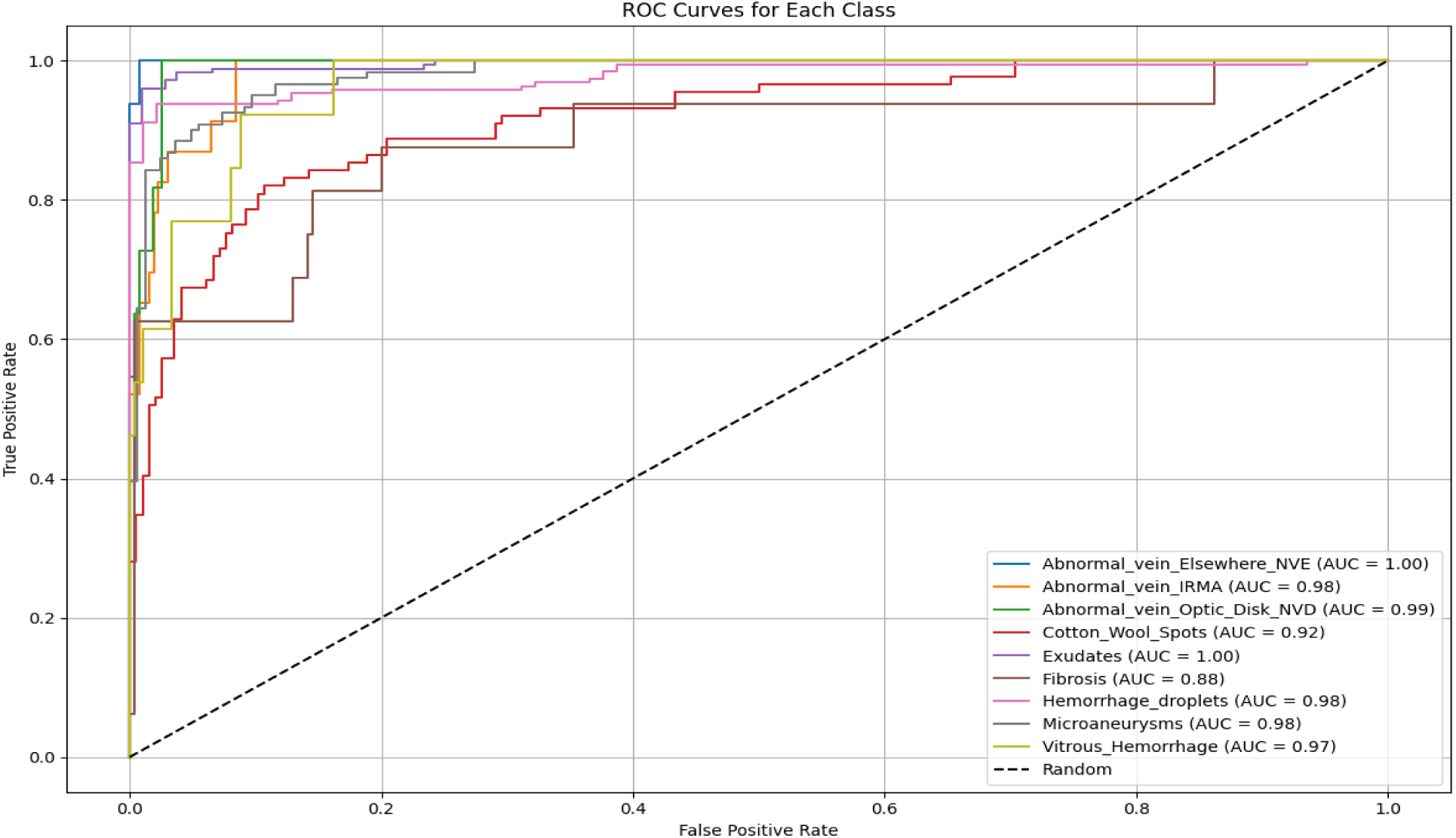
***ROC curves for aggregated clinical DR groupings.*** (A) Normal versus abnormal DR. (B) Non-referable versus referable DR. Near-optimal AUC values support the model’s clinical applicability for referral triage.

**Figure 8.**
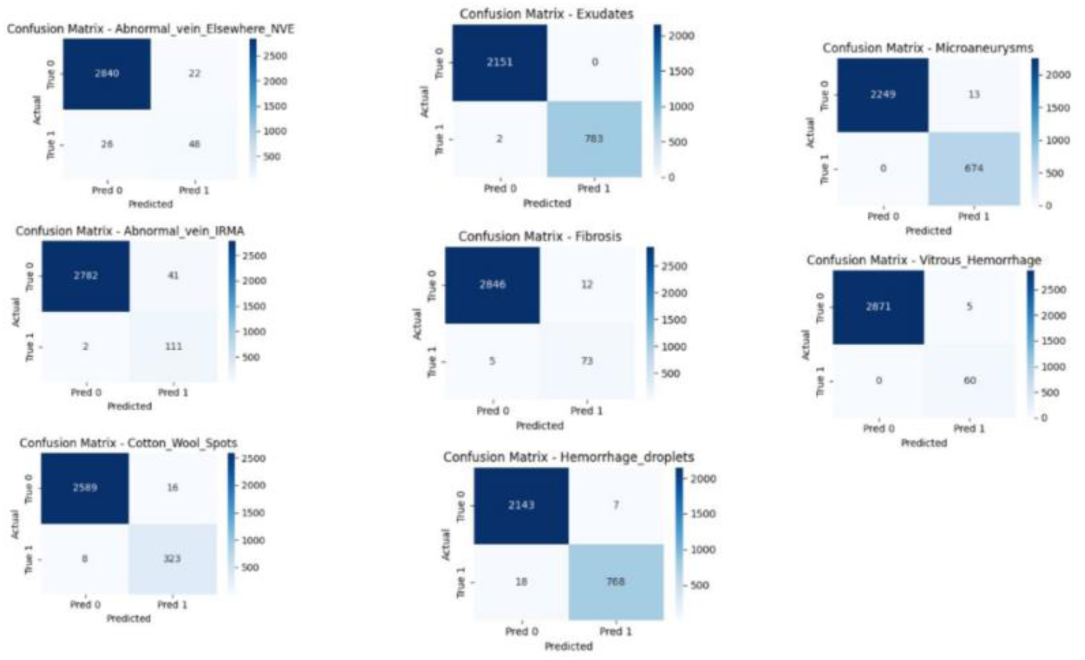
Confusion matrices for lesion-specific classification performance of the AI model. Confusion matrices are shown for detection of individual diabetic retinopathy lesions, including neovascularisation (abnormal vein/fibrovascular proliferation), exudates, microaneurysms, intraretinal microvascular abnormalities (IRMA), fibrosis, vitreous haemorrhage, cotton wool spots, and haemorrhagic droplets. Each matrix presents true versus predicted classifications, illustrating class-wise performance, with high true-positive rates and low misclassification across lesion categories, supporting robust lesion-level discrimination by the model.

### Multi-label lesion detection (Component II)

The companion multi-label lesion-detection head demonstrated strong per-lesion discriminatory performance (Table 4). Macro-averaged sensitivity was 93·9% (weighted: 97·9%), specificity was 99·5% (weighted: 99·6%), and AUC was 0·997 across nine lesion classes. Seven of nine classes achieved AUC = 1·00. NVD (AUC = 0·98) and NVE (AUC = 0·99) showed marginally lower performance consistent with their limited training sample sizes (n = 35 and n = 74, respectively). NVE recorded the lowest sensitivity (64·9%), attributable to class imbalance. Microaneurysms (sensitivity = 100·0%, AUC = 1·00) and exudates (sensitivity = 99·7%, AUC = 1·00) achieved near-perfect performance.

**Table 4.** Per-lesion classification performance of Component II on the held-out test set.

| Lesion | TP | FP | FN | TN | Sensitivity | Specificity | PPV | NPV | F1 | AUC |
| --- | --- | --- | --- | --- | --- | --- | --- | --- | --- | --- |
| NVE | 48 | 22 | 26 | 2840 | 64.9% | 99.2% | 68.6% | 99.1% | 0.667 | 0.99 |
| IRMA | 111 | 41 | 2 | 2782 | 98.2% | 98.6% | 73.0% | 99.9% | 0.838 | 1.00 |
| NVD | — | — | — | — | — | — | — | — | — | 0.98 |
| Cotton wool spots | 323 | 16 | 8 | 2589 | 97.6% | 99.4% | 95.3% | 99.7% | 0.964 | 1.00 |
| Exudates | 783 | 0 | 2 | 2151 | 99.7% | 100.0% | 100.0% | 99.9% | 0.999 | 1.00 |
| Fibrosis | 73 | 12 | 5 | 2846 | 93.6% | 99.6% | 85.9% | 99.8% | 0.896 | 1.00 |
| Haemorrhage droplets | 768 | 7 | 18 | 2143 | 97.7% | 99.7% | 99.1% | 99.2% | 0.984 | 1.00 |
| Microaneurysms | 674 | 13 | 0 | 2249 | 100.0% | 99.4% | 98.1% | 100.0% | 0.990 | 1.00 |
| Vitreous haemorrhage | 60 | 5 | 0 | 2871 | 100.0% | 99.8% | 92.3% | 100.0% | 0.960 | 1.00 |
| **Macro average** | — | — | — | — | **93.9%** | **99.5%** | — | — | **0.912** | **0.997** |

*AUC = area under the ROC curve (trapezoidal rule, one-versus-rest). NVD confusion matrix not generated owing to insufficient test-set samples (n ≈5); AUC derived from ROC curve legend. Macro average excludes NVD from sensitivity, specificity, PPV, NPV, and F1. PPV = positive predictive value; NPV = negative predictive value.*

### Lesion bounding-box detection output

Figures 10 and 11 illustrate the lesion-detection output of Component II applied to representative fundus images from the test set, displayed as bounding-box overlays derived from the pixel-level lesion masks. Figure 10 shows detections labelled by class index number (0–8), and Figure 11 shows the same detections with full class-name annotations. Red contours delineate vascular abnormalities (NVD, NVE); yellow bounding boxes denote individually detected lesion instances across all nine categories. The figures demonstrate the model’s capacity for simultaneous multi-lesion detection. Images 43346 and 43057 illustrate co-occurring NVD with microaneurysms and exudates, while images 46815 and 46573 show dense haemorrhage droplet clusters consistent with R2–M0 Severe DR.

**Figure 9.**
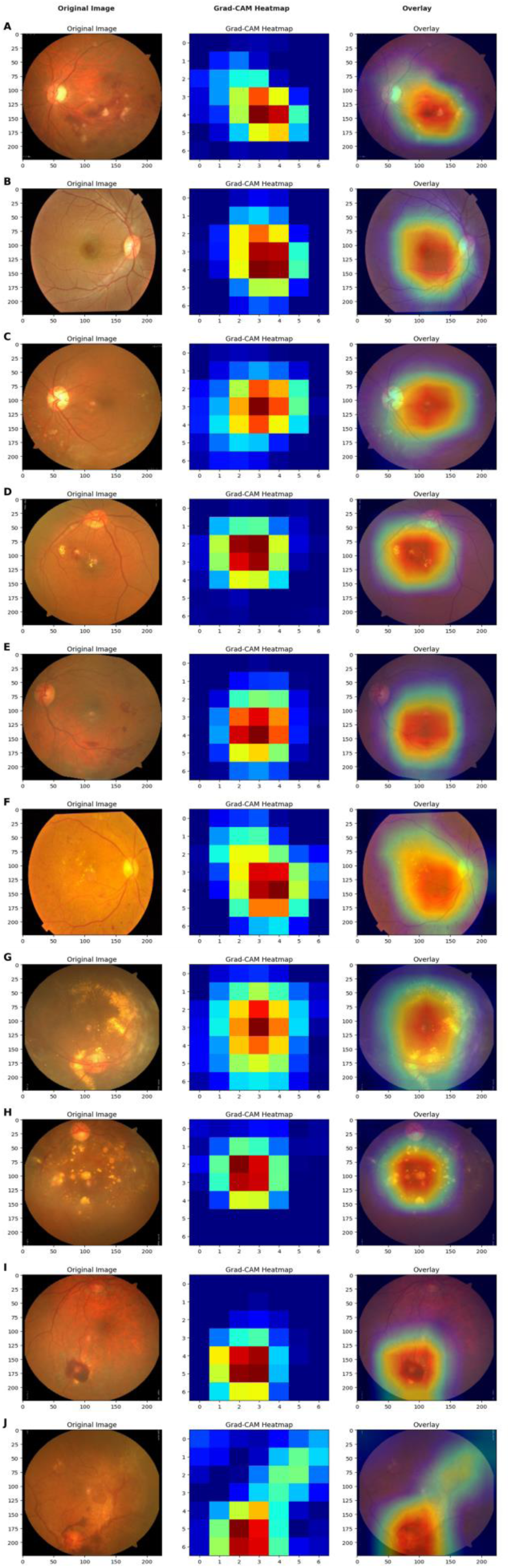
***Grad-CAM spatial attention heatmaps overlaid on representative fundus images across all ten UK DESP grades.*** Warmer colours (red–yellow) indicate higher model attention. (A, B) Non-DR and R0–M0: diffuse, low-intensity activation. (C, D) R1–M0 and R1–M1 mild DR: localised activation at early microaneurysms and exudates. (E–G) R2–M0 and R2–M1: broader activations covering haemorrhages and hard exudates. (H–J) R3–M0 and R3–M1 PDR: extensive peripheral activation consistent with neovascular and structural alterations.

**Figure 10.**
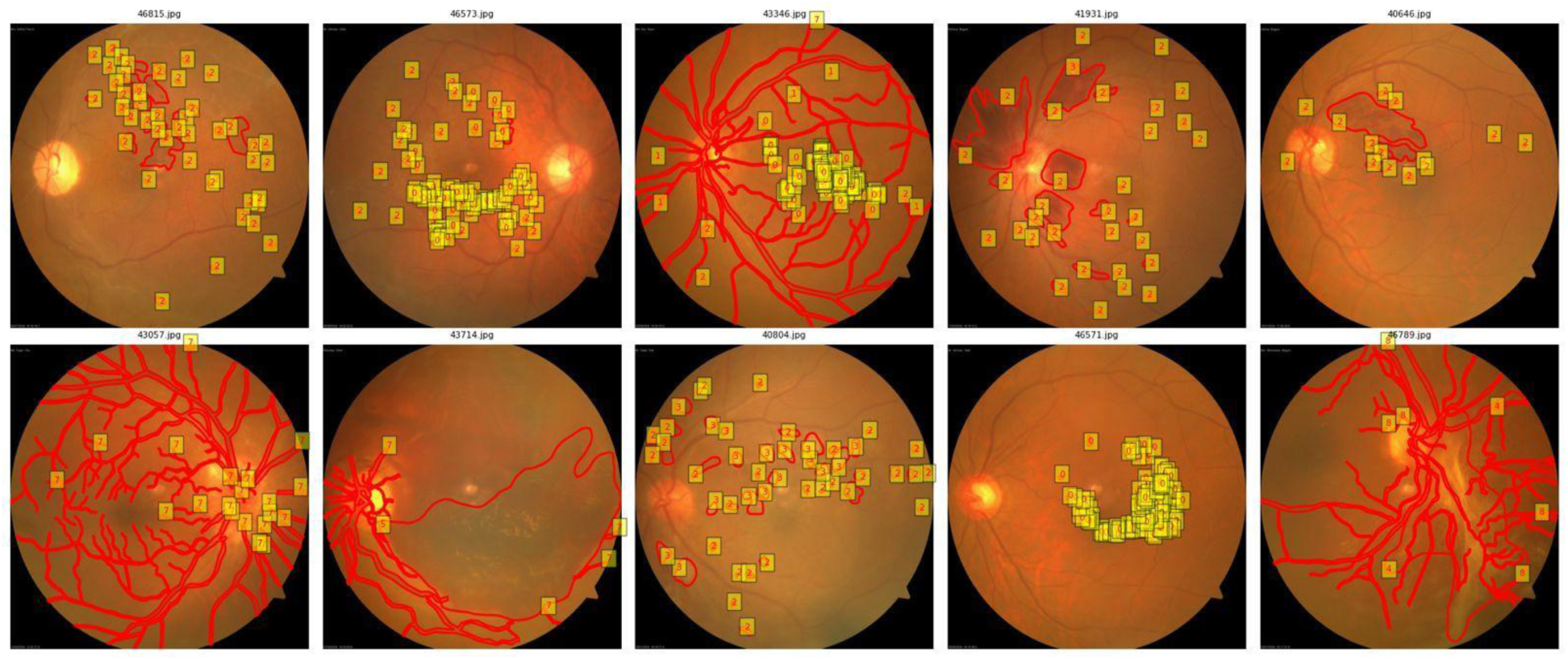
***Component II lesion-detection bounding-box output with class index labels on representative test-set fundus images.*** Yellow bounding boxes denote individually detected lesion instances; red contours delineate vascular abnormalities. Numbers within boxes indicate class index (0 = NVE; 1 = IRMA; 2 = NVD; 3 = Cotton wool spots; 4 = Exudates; 5 = Fibrosis; 6 = Haemorrhage droplets; 7 = Microaneurysms; 8 = Vitreous haemorrhage). Images 43057 and 43346 demonstrate dense NVD (class 2) co-occurring with microaneurysms, consistent with R3 proliferative DR; images 46815 and 46573 show dense haemorrhage droplet clusters (class 6) consistent with R2 Severe DR.

**Figure 11.**
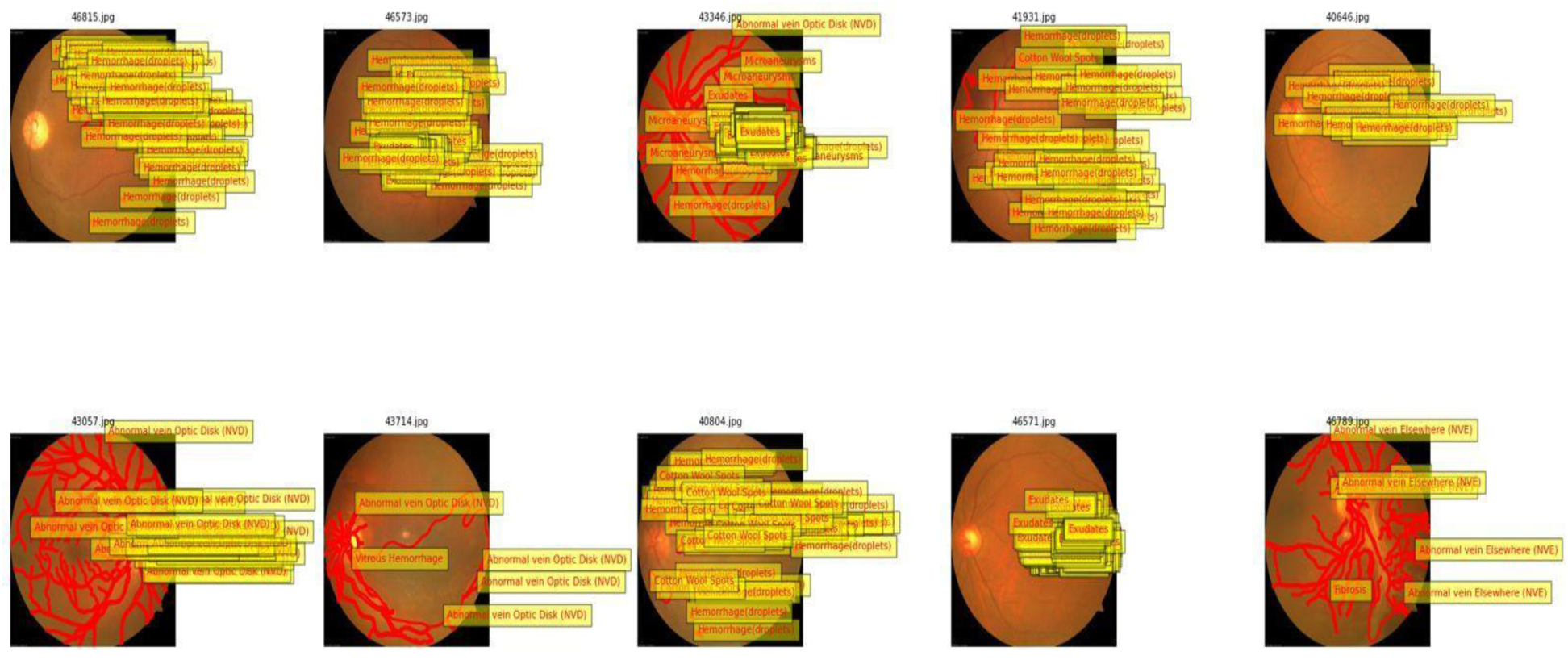
***Component II lesion-detection bounding-box output with full class-name labels on representative test-set fundus images.*** The same ten images as Figure 10 with full annotation names. Image 43714 demonstrates isolated vitreous haemorrhage with NVD (advanced proliferative DR); image 46571 shows macular exudate clustering consistent with M1 maculopathy; image 46789 demonstrates NVE with fibrosis consistent with R3 disease.

### Grad-CAM interpretability analysis

Grad-CAM activation maps (Figure 9) showed stage-dependent spatial attention patterns across all ten UK DESP classes. In non-DR and R0–M0 normal eyes, activations were diffuse without focal hotspots. In mild DR (R1–M0, R1–M1), attention localised to early microaneurysms and small exudative lesions. For moderate and severe DR (R2–M0, R2–M1), activations broadened to cover haemorrhages and hard exudates. In proliferative DR (R3–M0, R3–M1), activations extended toward lesion-dense peripheral areas. The high lesion-level AUCs (Table 4), particularly for microaneurysms and exudates (both AUCs = 1·00), corroborate the spatial fidelity of Grad-CAM activations in attending to pathologically relevant retinal regions. Mean inference time was 110–160 ms per image.

## Discussion

DRAGS demonstrates high diagnostic accuracy for ten-class UK DESP-aligned DR grading with clinically interpretable lesion-level explainability on a large real-world dataset from Bangladesh. To our knowledge, this is the first deep learning system to operationalise the full UK DESP taxonomy into a nine-class multi-label output integrated with a parallel lesion-detection head, spatial interpretability validation, and a pre-specified statistical analysis plan compliant with TRIPOD+AI. The overall accuracy of approximately 91·1%, quadratically weighted κ of approximately 0·90, and lesion-level macro-AUC of 0·997 compare favourably with published multi-class DR grading systems and with human inter-grader agreement benchmarks (κ ≈ 0·52).

The classification accuracy of DRAGS (91·1%) merits direct contextualisation against prior CNN-based approaches. Akhtar et al. reported 99·36% accuracy for their RSG-Net model on the Messidor-1 dataset comprising 1,200 retinal images.^6^ While this figure appears superior, it is attributable to a markedly smaller and less heterogeneous dataset. Models trained and evaluated on a few hundred to low thousands of images from a single source are susceptible to overfitting to dataset-specific characteristics. By contrast, DRAGS achieved 91·1% accuracy on a held-out test set of 3,044 images drawn from 20,281 fundus photographs acquired across two real-world Bangladeshi tertiary institutions with varying lighting conditions. Furthermore, RSG-Net performs only two- or four-class stratification without maculopathy subclassification, rendering its output unsuitable for direct UK DESP referral triage. DRAGS addresses precisely this limitation by integrating maculopathy status into a ten-class output taxonomy. This approach is clinically meaningful as M1 maculopathy elevates referral urgency due to its association with central vision risk.^20^

DRAGS similarly performs favourably against hybrid and multi-task architectures. Sheejakumari et al. reported 89·2% accuracy, 90·5% sensitivity, and 92% specificity for their MM-Net model on a dataset of only 3,663 images spanning five image categories.^23^ DRAGS surpasses MM-Net on all three metrics, achieving 91·1% accuracy, 90·7% sensitivity, and 91·9% specificity for referable versus non-referable DR, while classifying ten clinically interpretable UK DESP grades on a dataset nearly six times larger. Vijayalakshmi et al. proposed the MVCAViT architecture, integrating CNNs and Vision Transformers for multi-task DR grading, achieving attention-based lesion tracing on the DRTiD dataset.^22^ However, MVCAViT relies on bounding-box annotations to guide lesion localisation and cannot be readily generalised to datasets lacking such labels. DRAGS eludes this requirement: the grading component (Component I) requires no lesion-level annotation at inference time. At the same time, Grad-CAM overlays produce spatially faithful attention maps that are validated against pixel-level expert masks, providing interpretability without the annotation burden. Similarly, Abbasi et al. demonstrated improvements in sensitivity and specificity through adaptive gamma enhancement and histogram equalisation on a single dataset under controlled imaging conditions.^21^ The preprocessing pipeline adopted in DRAG, comprising Gaussian illumination correction and CLAHE, achieves comparable enhancement objectives while being validated across two institutions with differing acquisition devices, improving confidence in cross-device generalisability.

In the domain of lesion-level analysis, DRAGS builds upon and extends prior segmentation-grading paradigms. Porwal et al. demonstrated, using the IDRiD benchmark, that models combining lesion segmentation with image-level grading consistently outperform classification-only approaches in interpretability and clinical alignment. However, performance degrades when lesion annotations are skewed across DR severity grades.^24^ DRAGS directly instantiates this principle through its dual-component architecture: Component I assigns an overall DR severity grade, and Component II identifies the specific lesion features that underpin that grade, thereby providing a clinically auditable explanation for each prediction. The lesion-detection head achieved macro-averaged sensitivity of 93·9%, specificity of 99·5%, an AUC of 0·997 across nine lesion classes, with seven of nine classes reaching an AUC of 1·00. Guo and Peng’s CARNet demonstrated strong multi-lesion segmentation performance across three public datasets (IDRiD, E-Ophtha, DDR) using a cascade-attentive dual-encoder framework.^25^ However, CARNet focuses exclusively on pixel-level segmentation without incorporating disease severity grading, limiting its standalone clinical utility. DRAGS addresses this gap by integrating lesion detection and grading into a single end-to-end system. Incir and Bozkurt similarly proposed a two-stage joint segmentation-and-classification framework evaluated on the APTOS dataset, demonstrating that lesion-based weight maps improve severity grading over classification-only models.^27^ Their framework, however, requires fully annotated pixel-level lesion masks for training, raising scalability concerns in clinical settings where such annotations are rarely available. The ablation analysis in the present study confirmed that lesion-aware joint training in DRAGS similarly reduced confusion between adjacent severity grades (R2 versus R3) and maculopathy subgroups (M0 versus M1), consistent with the findings of Incir and Bozkurt, while the grading component does not depend on lesion masks at inference, a meaningful operational advantage for real-world deployment.^30^

Residual misclassification was predominantly confined to adjacent severity grades, mirroring the inter-observer variability inherent in fine-grained DR grading and consistent with off-diagonal error patterns observed in comparable multi-class systems. The low sensitivity for NVE (64·9%) reflects severe class imbalance (n = 74 training masks) and identifies neovascularisation categories as a priority for future dataset expansion. This limitation was also acknowledged in lesion-specific benchmarks such as IDRiD, where performance degradation under annotation imbalance was similarly documented.^24^ The model’s development and validation in a LMIC setting is a particular strength: over 80% of the global DR burden is concentrated in LMICs.^10^ Most published systems, including RSG-Net, MM-Net, MVCAViT, CARNet, and the IDRiD benchmarks, are validated exclusively on high-income-country or curated public datasets, limiting their applicability to the populations bearing the greatest disease burden.

Limitations include: (1) single-country dataset from two institutions, with performance potentially varying across populations, devices, and healthcare settings; (2) the companion lesion mask dataset is small (2,936 masks) with pronounced class imbalance (NVD n = 35, NVE n = 74), limiting generalisability of NVD/NVE performance estimates; (3) DenseNet-201’s computational requirements may constrain deployment on low-resource hardware, though its inference throughput (110–160 ms/image) supports teleophthalmology and PACS integration; and (4) Grad-CAM provides a class-discriminative but not causally faithful approximation of model attention.

## Conclusion

DRAGS is a dual-component deep learning framework that uniquely operationalises the UK DESP grading taxonomy into nine clinically interpretable classes, integrating classification with lesion-level detection and spatial interpretability. The system achieved approximately 91·1% accuracy, κ ≈ 0·90, and lesion-level macro-AUC of 0·997. Lesion-aware training improved discrimination at clinically challenging grade boundaries, and stage-dependent Grad-CAM patterns demonstrated biological plausibility of model attention. Prospective external validation is warranted before clinical deployment. In the future, we intend to combine image data with other clinical data, such as patient history and genetic information, to curate more precise and personalised diagnostic models.

## Data Availability

The fundus image dataset used in this study was acquired at Bangladesh Eye Hospital, Chattogram, Bangladesh, and cannot be publicly shared due to patient confidentiality restrictions and institutional data governance policies. Model code and weights are proprietary to Bangladesh Eye Hospital and are not available for external distribution. Researchers wishing to enquire about data or model access may contact the corresponding author. All analysis was conducted in accordance with TRIPOD+AI open science principles, and this data sharing statement is provided in fulfilment of those reporting requirements.

## Contributors

### Data sharing

The fundus image dataset used in this study was acquired at Bangladesh Eye Hospital Ltd., Dhaka, Bangladesh, and cannot be publicly shared due to patient confidentiality restrictions and institutional data governance policies. Model code and weights are proprietary to Bangladesh Eye Hospital Ltd. and are not available for external distribution. Researchers wishing to enquire about data or model access may contact the corresponding author. All analysis was conducted in accordance with TRIPOD+AI open science principles, and this data sharing statement is provided in fulfilment of those reporting requirements.

## References

1. Ong KL, Stafford LK, McLaughlin SA, et al. Global, regional, and national burden of diabetes from 1990 to 2021. Lancet 2023; 402: 203–34.

2. Kropp M, Golubnitschaja O, Mazurakova A, et al. Diabetic retinopathy as the leading cause of blindness and early predictor of cascading complications. EPMA J 2023; 14: 21–42.

3. Duh EJ, Sun JK, Stitt AW. Diabetic retinopathy: current understanding, mechanisms, and treatment strategies. JCI Insight 2017; 2: e93751.

4. Wahyu T, Syumarti. The epidemiology of diabetic retinopathy. EPMA J 2023; 14: 21–42.

5. Ghosh S, Chatterjee A. Transfer-ensemble learning based deep CNNs for diabetic retinopathy classification. 2019.

6. Akhtar S, Aftab S, Ali O, et al. A deep learning based model for diabetic retinopathy grading. Sci Rep 2025; 15: 1–20.

7. Abdelmaksoud E, El-Sappagh S, Barakat S, et al. Automatic diabetic retinopathy grading system based on detecting multiple retinal lesions. IEEE Access 2021; 9: 15939–60.

8. Li Z, Keel S, Liu C, et al. An automated grading system for detection of vision-threatening referable diabetic retinopathy. Diabetes Care 2018; 41: 2509–16.

9. Pao SI, Lin HZ, Chien KH, et al. Detection of diabetic retinopathy using bichannel CNN. J Ophthalmol 2020; 2020: 1972627.

10. Vujosevic S, Aldington SJ, Silva P, et al. Screening for diabetic retinopathy: new perspectives and challenges. Lancet Diabetes Endocrinol 2020; 8: 337–47.

11. Chan JCN, Lim LL, Wareham NJ, et al. The Lancet Commission on diabetes. Lancet 2020; 396: 2019–82.

12. Resnikoff S, Lansingh VC, Washburn L, et al. Estimated number of ophthalmologists worldwide. Br J Ophthalmol 2020; 104: 588–92.

13. Quinn L, Tryposkiadis K, Deeks JJ, et al. Interobserver variability studies in diagnostic imaging. Br J Radiol 2023; 96: 20220646.

14. Senapati A, Tripathy HK, Sharma V, Gandomi AH. AI for diabetic retinopathy detection: a systematic review. Inform Med Unlocked 2024; 45: 101445.

15. Rajesh AE, Davidson OQ, Lee CS, Lee AY. Artificial intelligence and diabetic retinopathy. Diabetes Care 2023; 46: 1728–39.

16. Wilkinson CP, Ferris FL 3rd, Klein RE, et al. Proposed international clinical diabetic retinopathy disease severity scales. Ophthalmology 2003; 110: 1677–82.

17. Bhaskaranand M, Ramachandra C, Bhat S, et al. The value of automated DR screening with EyeArt. Diabetes Technol Ther 2019; 21: 635–43.

18. Gulshan V, Peng L, Coram M, et al. Development and validation of a deep learning algorithm for DR detection. JAMA 2016; 316: 2402–10.

19. Takahashi H, Tampo H, Arai Y, et al. Applying AI to disease staging: deep learning for DR. PLoS One 2017; 12: e0179790.

20. NHS Diabetic Eye Screening Programme: grading definitions for referable disease. NHS England, 2025.

21. Abbasi R, Amin F, Alabrah A, et al. DR detection from fundus images using deep CNNs. Sci Rep 2025; 15: 6362–9.

22. Vijayalakshmi S, Manoharan JS, Nivetha B, Sathiya A. Multi-task deep learning framework combining CNN, vision transformers and PSO for DR diagnosis. Sci Rep 2025; 15: 35076.

23. Sheejakumari V, Sundravadivelu K, Pushparani S, et al. DR detection based on mobile maxout network. Sci Rep 2025; 15: 1–27.

24. Porwal P, Pachade S, Kokare M, et al. IDRiD: diabetic retinopathy — segmentation and grading challenge. Med Image Anal 2020; 59: 101561.

25. Guo Y, Peng Y. CARNet: cascade attentive RefineNet for multi-lesion segmentation. Complex Intell Syst 2022; 8: 1681–701.

26. Bidwai P, Gite S, Pradhan B, et al. Deep learning for detection of DR in geriatric group using OCTA. MethodsX 2024; 13: 102910.

27. Hattiya T, Dittakan K, Musikasuwan S. DR detection using CNN: a comparative study. Eng Access 2021; 7: 50–60.

28. Collins GS, Moons KGM, Dhiman P, et al. TRIPOD+AI statement: updated guidance for reporting clinical prediction models. BMJ 2024; 385: e078378.

29. Selvaraju RR, Cogswell M, Das A, et al. Grad-CAM: visual explanations from deep networks via gradient-based localisation. ICCV 2017; 618–26.

30. Incir R, Bozkurt F. A comprehensive deep-learning framework integrating lesion segmentation and stage classification for enhanced diabetic retinopathy diagnosis. Int J Imaging Syst Technol 2025; 35: e70272. 10.1002/ima.70272

